# Resources and Functional Maintenance Among Older Adults With Persistent Pain: A Harmonised Analysis of Three International Cohorts

**DOI:** 10.64898/2026.07.28.26359179

**Authors:** Lei Zhang, Wenjing Zhang, Linchao Li, Yuzhu Ye, Xiaona Zhu, Liang Shao, Huidi Yang, Yue Hu, Yuheng Li, Hai Lin, Wujun Geng

**Affiliations:** Department of Pain, The First Affiliated Hospital of Wenzhou Medical University, Wenzhou, Zhejiang, China; Department of Pain, Wenzhou Central Hospital, Affiliated to Wenzhou Medical University, Wenzhou, Zhejiang, China; Department of Anesthesia, The First Affiliated Hospital of Wenzhou Medical University, Wenzhou, Zhejiang, China; Department of Pain, Health Community Group of Yuhuan People’s Hospital, Yuhuan, Zhejiang, China; Wenzhou Medical University, Wenzhou, Zhejiang, China; Oujiang Laboratory (Zhejiang Lab for Regenerative Medicine, Vision and Brain Health), Wenzhou Medical University, Wenzhou, Zhejiang, China

**Keywords:** persistent pain, activities of daily living, physical activity, functional maintenance, ageing

## Abstract

**Background:** Persistent pain in later life is associated with functional decline. Identifying resources associated with preserved daily function may broaden pain research beyond symptom burden alone.

**Objective:** To examine six prespecified resource indicators—physical activity, depressive symptoms, recall performance, current partner status, education and wealth—in relation to subsequent maintenance of independence in activities of daily living (ADL).

**Methods:** This longitudinal observational study analysed adults aged 50 years or older with pain at both the initial and baseline assessments in the Health and Retirement Study (HRS), the English Longitudinal Study of Ageing (ELSA) and the Survey of Health, Ageing and Retirement in Europe (SHARE). ADL maintenance was defined as no difficulty in all five prespecified ADL activities at baseline and no difficulty in all five at follow-up. Indicator-specific risk ratios were estimated using modified Poisson regression with robust variance and pooled using random-effects meta-analysis. The six meta-analysis *P* values were adjusted using the Holm procedure. Each indicator used a separate complete-case sample; no common six-indicators sample was constructed.

**Results:** The descriptive samples comprised 8,279 HRS person-windows contributed by 4,688 unique participants, 1,200 ELSA participants and 6,995 SHARE participants; indicator-specific denominators varied. Physical activity was associated with a higher probability of ADL maintenance across all three cohorts (pooled risk ratio, 1.145; 95% CI, 1.097–1.195; *P* = .005; Holm-adjusted *P* = .032) and was the only prespecified hypothesis to meet the Holm-adjusted criterion. The other five hypotheses did not meet this criterion; failure to do so should not be interpreted as evidence that the corresponding associations were absent. Several of these pooled estimates showed substantial heterogeneity.

**Conclusions:** Among older adults with persistent pain, physical activity was associated with a higher probability of ADL maintenance, and the physical activity hypothesis was the only one of the six prespecified hypotheses to meet the Holm-adjusted criterion in the three-cohort meta-analysis. These observational findings support further study of physical activity and functional maintenance but do not establish causal benefit.

**Key Points:**

- Three international ageing cohorts were harmonised to evaluate associations between six resource-related indicators and subsequent ADL maintenance among older adults with persistent pain.
- Physical activity was consistently associated with a higher probability of ADL maintenance and was the only one of the six prespecified hypotheses to meet the Holm-adjusted criterion.
- The other five hypotheses did not meet the prespecified Holm-adjusted criterion, and several pooled estimates showed substantial heterogeneity.
- The findings are observational and do not establish that increasing physical activity would preserve ADL independence.

## Introduction

Persistent pain in later life is associated with functional limitations and decline [1–4]. Independence in dressing, bathing, eating, transferring and toileting is important to healthy ageing [2]. Functional trajectories vary: some older adults remain independent despite persistent pain, whereas others develop limitations [5].

This variation supports examining measurable characteristics associated with maintaining independence rather than focusing on pain burden and vulnerability alone. The six prespecified indicators represented behavioural, psychological, cognitive, social and socioeconomic domains.

Physical activity represented the behavioural domain and has been linked to pain-related or functional outcomes [6–8]; depressive symptoms and recall performance represented psychological and cognitive function with established links to disability or ADL [9,10]; current partner status represented social context, informed by broader evidence on social vulnerability and well-being [11–14]; and education and wealth represented socioeconomic position associated with disability and healthy ageing [15–17].

Existing longitudinal evidence has primarily characterised pain-related functional decline [3,4]; to our knowledge, the comparative associations of these six indicators with ADL maintenance have not been examined across harmonised international cohorts of adults with persistent pain. We therefore examined the six indicators in HRS, ELSA and SHARE, estimated cohort-specific associations, and pooled compatible estimates using random-effects meta-analysis. The primary objective was to identify which associations met the prespecified Holm-adjusted criterion, interpreted alongside cohort-specific estimates and between-cohort heterogeneity.

## Methods

### Study design

We conducted a longitudinal observational study using three international ageing cohorts: the Health and Retirement Study (HRS), the English Longitudinal Study of Ageing (ELSA) and the Survey of Health, Ageing and Retirement in Europe (SHARE). The study population comprised adults aged 50 years or older with persistent pain, defined as pain reported at both the initial assessment (T0) and the baseline assessment (T1). Resource indicators and baseline activities of daily living (ADL) were measured at T1, and ADL maintenance was assessed at follow-up (T2). The interval between T0 and T1 was approximately two years, whereas follow-up duration from T1 to T2 was fixed at exactly four years in the frozen analysis data for all three primary cohorts. The China Health and Retirement Longitudinal Study (CHARLS) was reserved for an extended sensitivity role because its pain-confirmation interval was not fully comparable with those of HRS, ELSA and SHARE; it was not included in the primary analysis. Cohort-specific assessment sequences and harmonisation procedures are detailed in Appendix S1 and Tables S1–S4.

### Cohorts

#### HRS

The HRS is a nationally representative US longitudinal study of ageing that collects multidisciplinary data [18]. The analysis included all prespecified eligible analysis windows before the COVID-19 pandemic, with each eligible window treated as one person-window. Participants could contribute more than one eligible person-window. The HRS estimate therefore represents the average association across eligible person-windows rather than a person-level association based on one selected observation per participant. Models used robust sandwich variance clustered by participant to account for repeated observations and included fixed effects for analysis window.

### ELSA

ELSA is a representative longitudinal study of ageing in England designed to be comparable with HRS [19]. The analysis used a single prespecified analysis window, with each participant contributing one observation. Models used heteroscedasticity-robust sandwich variance.

### SHARE

SHARE is a multinational, multidisciplinary panel study designed to support cross-national comparisons and harmonisation with HRS and ELSA [20]. The SHARE analysis used the prespecified Wave 4–Wave 5–Wave 7 window, with each participant contributing one observation. Models included fixed effects for baseline country and used heteroscedasticity-robust sandwich variance.

### Ethics

The original HRS, ELSA and SHARE studies were conducted under approvals from their respective ethics committees, and participants provided written informed consent in accordance with cohort-specific procedures. This secondary analysis used de-identified data obtained through the official access procedures of the respective cohorts. The relevant institutional body at the First Affiliated Hospital of Wenzhou Medical University determined that no additional ethics approval was required for this secondary analysis; no reference number was issued.

### Participants

Eligible observations required successful linkage between T0 and T1, valid pain information and reported pain at both assessments. Eligibility further required age 50 years or older at T1, a valid value for the indicator under analysis and successful linkage to T2 under the prespecified rules for outcome observation. Observations were restricted to those with no difficulty in all five ADL activities at T1 and valid information for all five activities at T2.

Each indicator was analysed in a separate complete-case sample; no common complete-case sample across all six indicators was constructed. Detailed sample flow, indicator-specific sample sizes and outcome counts are provided in Appendix S3 and Tables S5–S6.

### Exposure definitions

The six prespecified resource-related indicators were physical activity, depressive symptoms, recall performance, current partner status, education and wealth. Physical activity was modelled as active versus inactive, and current partner status as having versus not having a current partner. Depressive-symptom and recall scores were standardised separately within each cohort and assessment wave and were modelled per 1-SD increase; higher scores indicated more depressive symptoms and better recall, respectively. Education was modelled as an ordinal trend across low, medium and high categories, and wealth as an ordinal trend across quintiles, with risk ratios expressed per one-category and one-quintile increase, respectively. Prespecified indicator categories and coding rules were retained without alteration.

### Outcome

The primary outcome was ADL maintenance. Participants were included in the ADL-maintenance analysis if they reported no difficulty in any of the five prespecified ADL activities at T1. Among those with valid information for all five activities at T2, ADL maintenance was defined as remaining free of difficulty in all five. Death, non-death loss to follow-up, refusal, non-entry into the relevant module and structural missingness were distinguished from observed ADL outcomes and were not coded as failure to maintain ADL.

### Covariates

All cohort-specific models adjusted for sex and for age modelled using a prespecified restricted cubic spline with three knots. HRS models included analysis-window fixed effects and used participant-clustered robust sandwich variance, while SHARE models included fixed effects for baseline country. Follow-up duration from T1 to T2 was fixed at exactly four years for every observation in the frozen analysis data for all three primary cohorts. It was therefore exactly collinear with the intercept and was omitted from the final models because it provided no independently estimable information. The corresponding design-matrix audit and verification are reported in Appendix S5 and Table S8.

### Statistical analysis

For each indicator within each cohort, we fitted a separate modified Poisson regression model with a log link to estimate risk ratios [21]. RRs were estimated from the fitted models, while 95% CIs and two-sided P values were based on robust sandwich variance estimates; inference therefore did not require the Poisson mean–variance equality assumption. HRS used robust sandwich variance clustered by participant to account for repeated person-windows, whereas ELSA and SHARE used heteroscedasticity-robust sandwich variance. The primary analyses did not apply sampling weights and used separate complete-case samples for each indicator.

For each indicator, the corresponding cohort-specific log RRs and standard errors from HRS, ELSA and SHARE were combined in a separate random-effects meta-analysis. Between-cohort variance (τ²) was estimated by restricted maximum likelihood (REML), and 95% CIs were based on the Hartung–Knapp method [22]. Pooled estimates and their confidence limits were transformed back to the RR scale. We reported τ², I² and Cochran’s Q. Because only three cohorts contributed to each meta-analysis, heterogeneity statistics were interpreted cautiously [23]. Hartung–Knapp confidence intervals were calculated using a t distribution with two degrees of freedom [22] and were likewise interpreted cautiously.

The confirmatory hypothesis family comprised the six prespecified ADL-maintenance hypotheses evaluated at the three-cohort meta-analysis level. The Holm step-down procedure was used to control the family-wise error rate across this six-hypothesis family [24]. The two-sided family-wise significance level was 0.05. Both unadjusted and Holm-adjusted P values were reported. Only a Holm-adjusted P value below 0.05 was considered to meet the prespecified confirmatory multiplicity criterion. Sampling-weighted analyses, inverse-probability weighting for T2 retention and multiple imputation were not performed.

## Results

### Cohort and sample description

The descriptive samples comprised 8,279 HRS person-windows contributed by 4,688 unique participants, 1,200 ELSA participants and 6,995 SHARE participants. Cohort-specific sample formation and indicator-specific sample sizes are shown in Figure 1. Mean age was 66.70 years (SD, 9.23) in HRS, 66.09 years (SD, 8.56) in ELSA and 67.16 years (SD, 9.01) in SHARE. Women accounted for 5,389 person-windows (65.1%) in HRS, 773 participants (64.4%) in ELSA and 4,630 participants (66.2%) in SHARE. Table 1 presents ADL independence, the six resource-related indicators, BMI, multimorbidity, current smoking and current alcohol use, using valid denominators specific to each variable. BMI was unavailable in ELSA under the prespecified same-wave rule, and values from other waves were not used.

**Figure 1.**
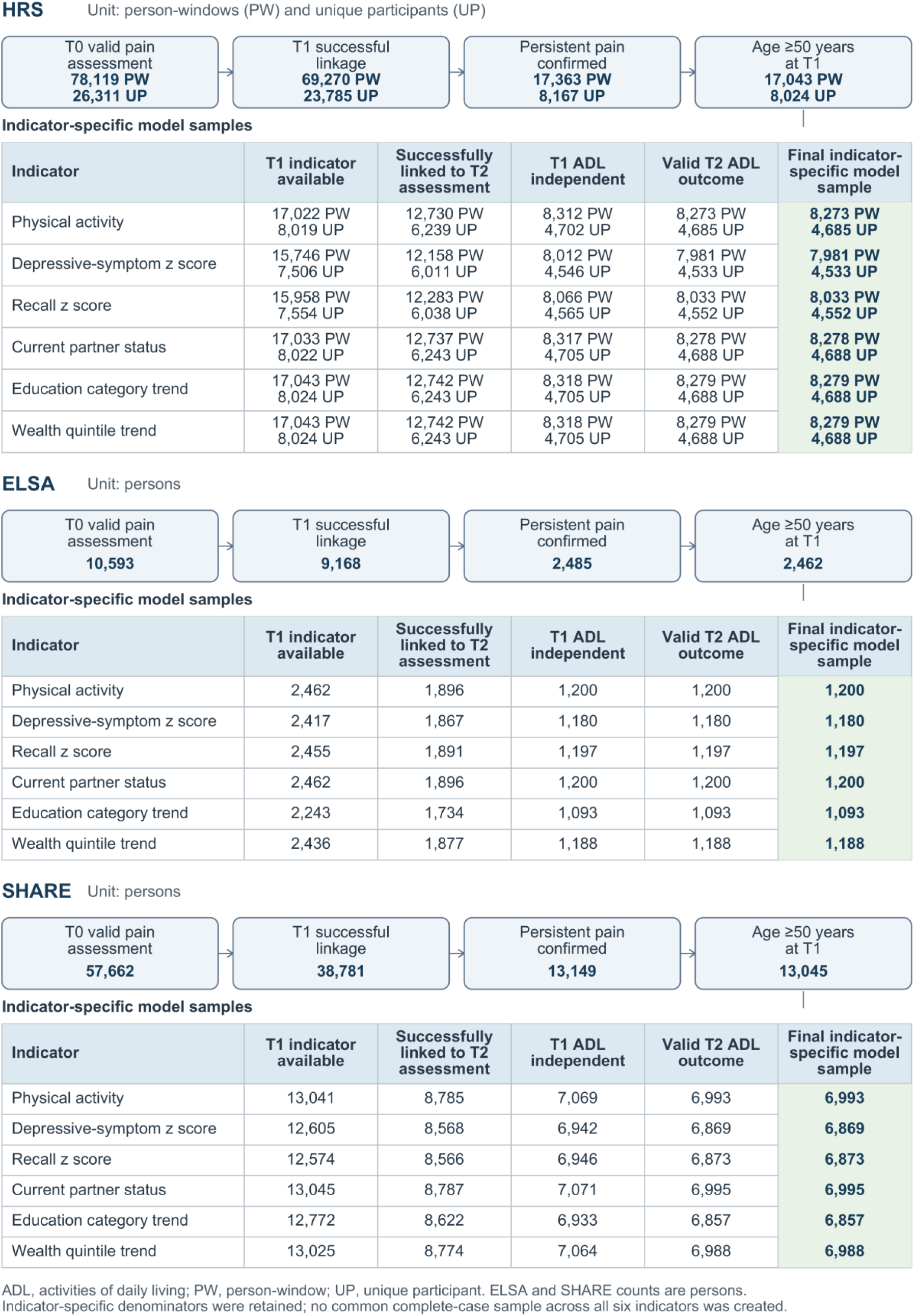
Formation of the primary activities of daily living analysis samples. The figure presents the prespecified cumulative display of sample formation in the Health and Retirement Study (HRS), English Longitudinal Study of Ageing (ELSA) and Survey of Health, Ageing and Retirement in Europe (SHARE). HRS counts are reported as person-windows and corresponding unique participants, whereas ELSA and SHARE counts are reported as participants. Within each indicator-specific branch, the displayed order comprises T1 indicator availability, successful linkage to the T2 assessment, T1 ADL independence, valid T2 ADL information and the final model sample. This order describes intermediate sample losses and does not represent the execution order of the primary model code; final samples were defined by the joint intersection of all applicable conditions. Indicator-specific denominators were retained, and no common complete-case sample across all six indicators was constructed. ADL indicates activities of daily living; PW, person-window; UP, unique participant. Alt text: Three-column flow diagram showing formation of the primary ADL analysis samples in HRS, ELSA, and SHARE from valid pain assessment through persistent-pain confirmation, age and indicator eligibility, follow-up outcome observation, baseline ADL independence, and resource-specific final samples. HRS displays person-window and unique-participant counts separately; ELSA and SHARE display participant counts. Six resource-specific denominators are shown rather than one common complete-case denominator.

**Table 1.** Descriptive Characteristics of the ADL-Maintenance Samples in HRS, ELSA and SHARE. Characteristics are presented for the frozen primary ADL analysis samples. HRS is reported using person-windows as the primary analysis unit, with unique participants also reported. ELSA and SHARE are reported at the person level. Continuous variables are presented as mean (SD), and categorical variables as number (percentage), using variable-specific valid denominators.

| Characteristic | HRS value | Valid n | Missing | ELSA value | Valid n | Missing | SHARE value | Valid n | Missing |
| --- | --- | --- | --- | --- | --- | --- | --- | --- | --- |
| Analysis sample, n | 8279 | 8279 | 0 | 1200 | 1200 | 0 | 6995 | 6995 | 0 |
| Unique participants, n | 4688 | — | — | 1200 | — | — | 6995 | — | — |
| Age, years, mean (SD) | 66.70 (9.23) | 8279 | 0 | 66.09 (8.56) | 1200 | 0 | 67.16 (9.01) | 6995 | 0 |
| Sex:Female | 5389 (65.1%) | 8279 | 0 | 773 (64.4%) | 1200 | 0 | 4630 (66.2%) | 6995 | 0 |
| Sex:Male | 2890 (34.9%) | 8279 | 0 | 427 (35.6%) | 1200 | 0 | 2365 (33.8%) | 6995 | 0 |
| Persistent pain, n (%) | 8279 (100.0%) | 8279 | 0 | 1200 (100.0%) | 1200 | 0 | 6995 (100.0%) | 6995 | 0 |
| ADL independent at baseline, n (%) | 8279 (100.0%) | 8279 | 0 | 1200 (100.0%) | 1200 | 0 | 6995 (100.0%) | 6995 | 0 |
| Physically active, n (%) | 5363 (64.8%) | 8273 | 6 | 911 (75.9%) | 1200 | 0 | 5903 (84.4%) | 6993 | 2 |
| Standardised depressive-symptom score, mean (SD) | 0.20 (1.06) | 7981 | 298 | 0.22 (1.07) | 1180 | 20 | 0.23 (1.00) | 6869 | 126 |
| Standardised recall score, mean (SD) | 0.12 (0.91) | 8033 | 246 | 0.06 (0.96) | 1197 | 3 | 0.03 (0.96) | 6873 | 122 |
| Current partner, n (%) | 5381 (65.0%) | 8278 | 1 | 890 (74.2%) | 1200 | 0 | 4806 (68.7%) | 6995 | 0 |
| Education:Low | 1475 (17.8%) | 8279 | 0 | 439 (40.2%) | 1093 | 107 | 3094 (45.1%) | 6857 | 138 |
| Education:Medium | 3111 (37.6%) | 8279 | 0 | 322 (29.5%) | 1093 | 107 | 2574 (37.5%) | 6857 | 138 |
| Education:High | 3693 (44.6%) | 8279 | 0 | 332 (30.4%) | 1093 | 107 | 1189 (17.3%) | 6857 | 138 |
| Wealth: Quintile 1 | 1597 (19.3%) | 8279 | 0 | 262 (22.1%) | 1188 | 12 | 1281 (18.3%) | 6988 | 7 |
| Wealth: Quintile 2 | 1845 (22.3%) | 8279 | 0 | 236 (19.9%) | 1188 | 12 | 1453 (20.8%) | 6988 | 7 |
| Wealth: Quintile 3 | 1749 (21.1%) | 8279 | 0 | 254 (21.4%) | 1188 | 12 | 1550 (22.2%) | 6988 | 7 |
| Wealth: Quintile 4 | 1610 (19.4%) | 8279 | 0 | 220 (18.5%) | 1188 | 12 | 1380 (19.7%) | 6988 | 7 |
| Wealth: Quintile 5 | 1478 (17.9%) | 8279 | 0 | 216 (18.2%) | 1188 | 12 | 1324 (18.9%) | 6988 | 7 |
| BMI, kg/m<sup>2</sup>, mean (SD) | 29.65 (6.39) | 8176 | 103 | NA | 0 | 1200 | 27.82 (5.09) | 6818 | 177 |
| Common chronic-condition count, mean (SD) | 2.59 (1.36) | 8279 | 0 | 1.62 (1.13) | 1200 | 0 | 1.44 (1.10) | 6993 | 2 |
| Multimorbidity ($\geq 2$ conditions), n (%) | 6558 (79.2%) | 8279 | 0 | 604 (50.3%) | 1200 | 0 | 3056 (43.7%) | 6993 | 2 |
| Current smoking, n (%) | 1193 (14.5%) | 8231 | 48 | 172 (14.3%) | 1200 | 0 | 1193 (17.1%) | 6973 | 22 |
| Current alcohol use, n (%) | 4272 (51.6%) | 8279 | 0 | 966 (86.4%) | 1118 | 82 | 4619 (66.1%) | 6992 | 3 |

1. ADL indicates activities of daily living; HRS, Health and Retirement Study; ELSA, English Longitudinal Study of Ageing; SHARE, Survey of Health, Ageing and Retirement in Europe. 2. HRS person-window counts are not counts of mutually independent participants; the unique participant count is reported separately. 3. Descriptive variables were summarised using their own valid denominators and did not redefine the indicator-specific primary model samples. 4. BMI uses official harmonised BMI from the same T1 wave. ELSA BMI is NA because no eligible same-wave harmonised BMI was available and values were not supplemented from another wave. 5. Multimorbidity is defined as at least two conditions across the eight approved common disease domains: hypertension, diabetes, heart disease, stroke, chronic lung disease, cancer, arthritis or rheumatism, and mental or emotional disease. 6. Smoking is presented as current versus not current. 7. Alcohol definitions were harmonised as current drinking status; recall periods differed across cohorts. 8. Table 1 is descriptive and was not used to redefine the primary model samples, meta-analysis, or Holm family.

### Cohort-specific associations

Table 2 presents the cohort-specific associations. Physical activity was associated with a higher probability of ADL maintenance in all three cohorts: HRS (RR, 1.139; 95% CI, 1.109– 1.170), ELSA (RR, 1.209; 95% CI, 1.111–1.315) and SHARE (RR, 1.144; 95% CI, 1.102–1.186). Higher depressive-symptom scores were associated with a lower probability of ADL maintenance in all three cohorts. The RR per 1-SD increase was 0.931 (95% CI, 0.919– 0.944) in HRS, 0.916 (95% CI, 0.887–0.946) in ELSA and 0.960 (95% CI, 0.950–0.971) in SHARE.

**Table 2.**
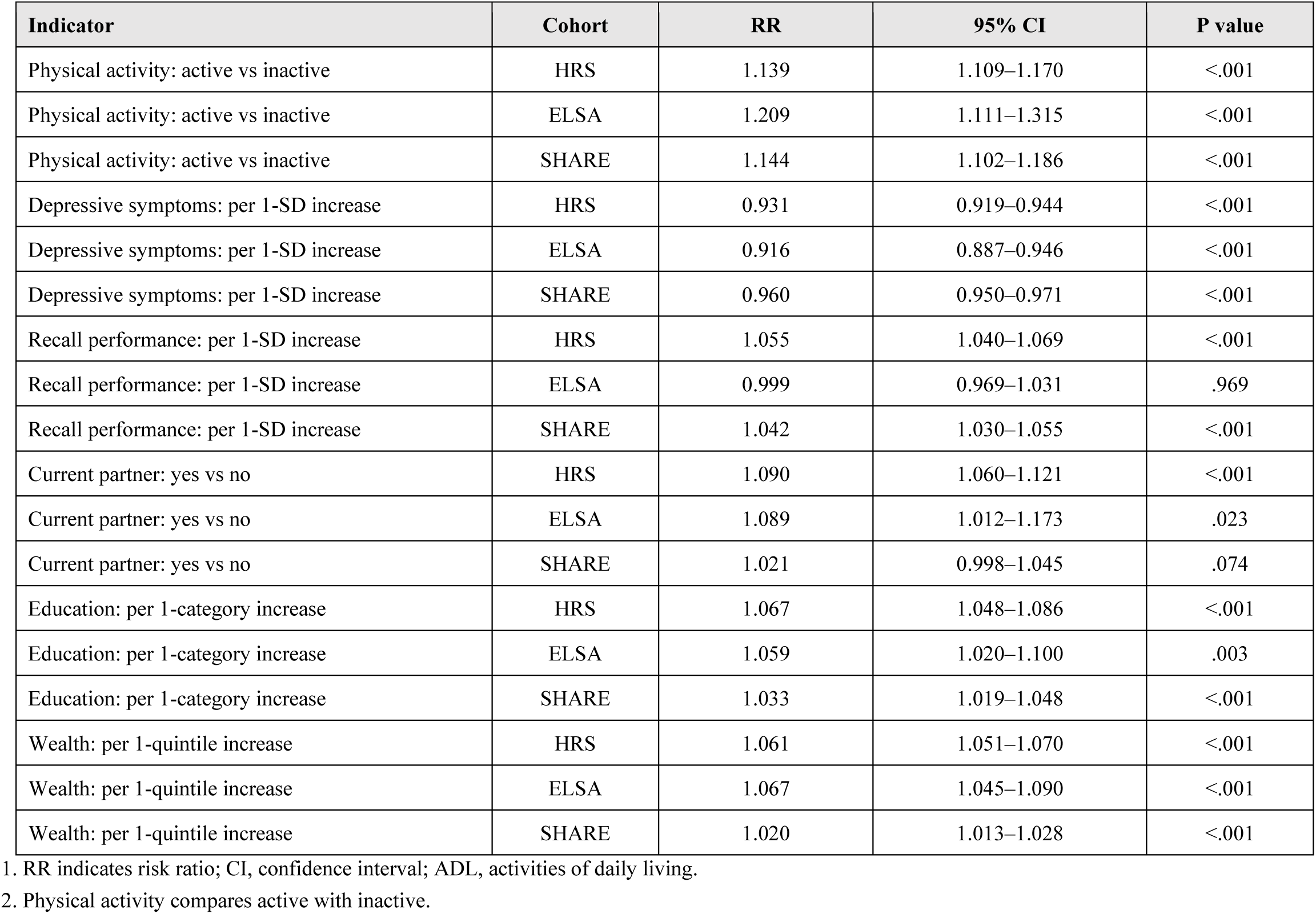
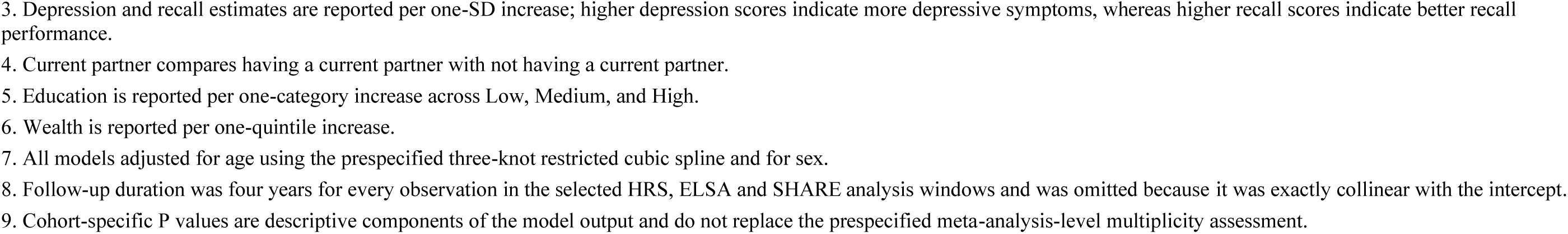
Cohort-Specific Associations Between Six Prespecified Resource-Related Indicators and ADL Maintenance. Risk ratios and 95% confidence intervals are from indicator-specific modified Poisson regression models with robust sandwich variance. Each indicator used its own complete-case sample. HRS models used all eligible pre-pandemic person-windows, participant-level clustering, and window fixed effects. ELSA models used person-level robust variance. SHARE models used person-level robust variance and country fixed effects.

| Indicator | Cohort | RR | 95% CI | P value |
| --- | --- | --- | --- | --- |
| Physical activity: active vs inactive | HRS | 1.139 | 1.109–1.170 | <.001 |
| Physical activity: active vs inactive | ELSA | 1.209 | 1.111–1.315 | <.001 |
| Physical activity: active vs inactive | SHARE | 1.144 | 1.102–1.186 | <.001 |
| Depressive symptoms: per 1-SD increase | HRS | 0.931 | 0.919–0.944 | <.001 |
| Depressive symptoms: per 1-SD increase | ELSA | 0.916 | 0.887–0.946 | <.001 |
| Depressive symptoms: per 1-SD increase | SHARE | 0.960 | 0.950–0.971 | <.001 |
| Recall performance: per 1-SD increase | HRS | 1.055 | 1.040–1.069 | <.001 |
| Recall performance: per 1-SD increase | ELSA | 0.999 | 0.969–1.031 | .969 |
| Recall performance: per 1-SD increase | SHARE | 1.042 | 1.030–1.055 | <.001 |
| Current partner: yes vs no | HRS | 1.090 | 1.060–1.121 | <.001 |
| Current partner: yes vs no | ELSA | 1.089 | 1.012–1.173 | .023 |
| Current partner: yes vs no | SHARE | 1.021 | 0.998–1.045 | .074 |
| Education: per 1-category increase | HRS | 1.067 | 1.048–1.086 | <.001 |
| Education: per 1-category increase | ELSA | 1.059 | 1.020–1.100 | .003 |
| Education: per 1-category increase | SHARE | 1.033 | 1.019–1.048 | <.001 |
| Wealth: per 1-quintile increase | HRS | 1.061 | 1.051–1.070 | <.001 |
| Wealth: per 1-quintile increase | ELSA | 1.067 | 1.045–1.090 | <.001 |
| Wealth: per 1-quintile increase | SHARE | 1.020 | 1.013–1.028 | <.001 |

1. RR indicates risk ratio; CI, confidence interval; ADL, activities of daily living.
2. Physical activity compares active with inactive.
3. Depression and recall estimates are reported per one-SD increase; higher depression scores indicate more depressive symptoms, whereas higher recall scores indicate better recall performance. 4. Current partner compares having a current partner with not having a current partner. 5. Education is reported per one-category increase across Low, Medium, and High. 6. Wealth is reported per one-quintile increase. 7. All models adjusted for age using the prespecified three-knot restricted cubic spline and for sex. 8. Follow-up duration was four years for every observation in the selected HRS, ELSA and SHARE analysis windows and was omitted because it was exactly collinear with the intercept. 9. Cohort-specific P values are descriptive components of the model output and do not replace the prespecified meta-analysis-level multiplicity assessment.

Recall performance showed greater cross-cohort variation. The RR per 1-SD increase in the standardised recall score was 1.055 (95% CI, 1.040–1.069) in HRS and 1.042 (95% CI, 1.030–1.055) in SHARE, whereas the ELSA estimate was close to 1.00 (RR, 0.999; 95% CI, 0.969–1.031). Estimates for having versus not having a current partner were above 1.00 in all three cohorts but were smaller in SHARE: 1.090 (95% CI, 1.060–1.121) in HRS, 1.089 (95% CI, 1.012–1.173) in ELSA and 1.021 (95% CI, 0.998–1.045) in SHARE.

Education and wealth estimates were above 1.00 in all three cohorts, although their magnitudes varied. For each one-category increase in education, the RR was 1.067 (95% CI, 1.048–1.086) in HRS, 1.059 (95% CI, 1.020–1.100) in ELSA and 1.033 (95% CI, 1.019–1.048) in SHARE. For each one-quintile increase in wealth, the RR was 1.061 (95% CI, 1.051–1.070) in HRS, 1.067 (95% CI, 1.045–1.090) in ELSA and 1.020 (95% CI, 1.013–1.028) in SHARE.

### Meta-analysis and multiplicity correction

Table 3 presents the three-cohort meta-analysis estimates and Holm-adjusted P values, and Figure 2 shows the cohort-specific and pooled estimates for all six indicators. Physical activity was associated with a higher probability of ADL maintenance (pooled RR, 1.145; 95% CI, 1.097–1.195; *P* = .005; Holm-adjusted *P* = .032). Estimated between-cohort heterogeneity was minimal (I² = 0.6%). The physical activity hypothesis was the only one of the six prespecified hypotheses to meet the Holm-adjusted criterion.

**Figure 2.**
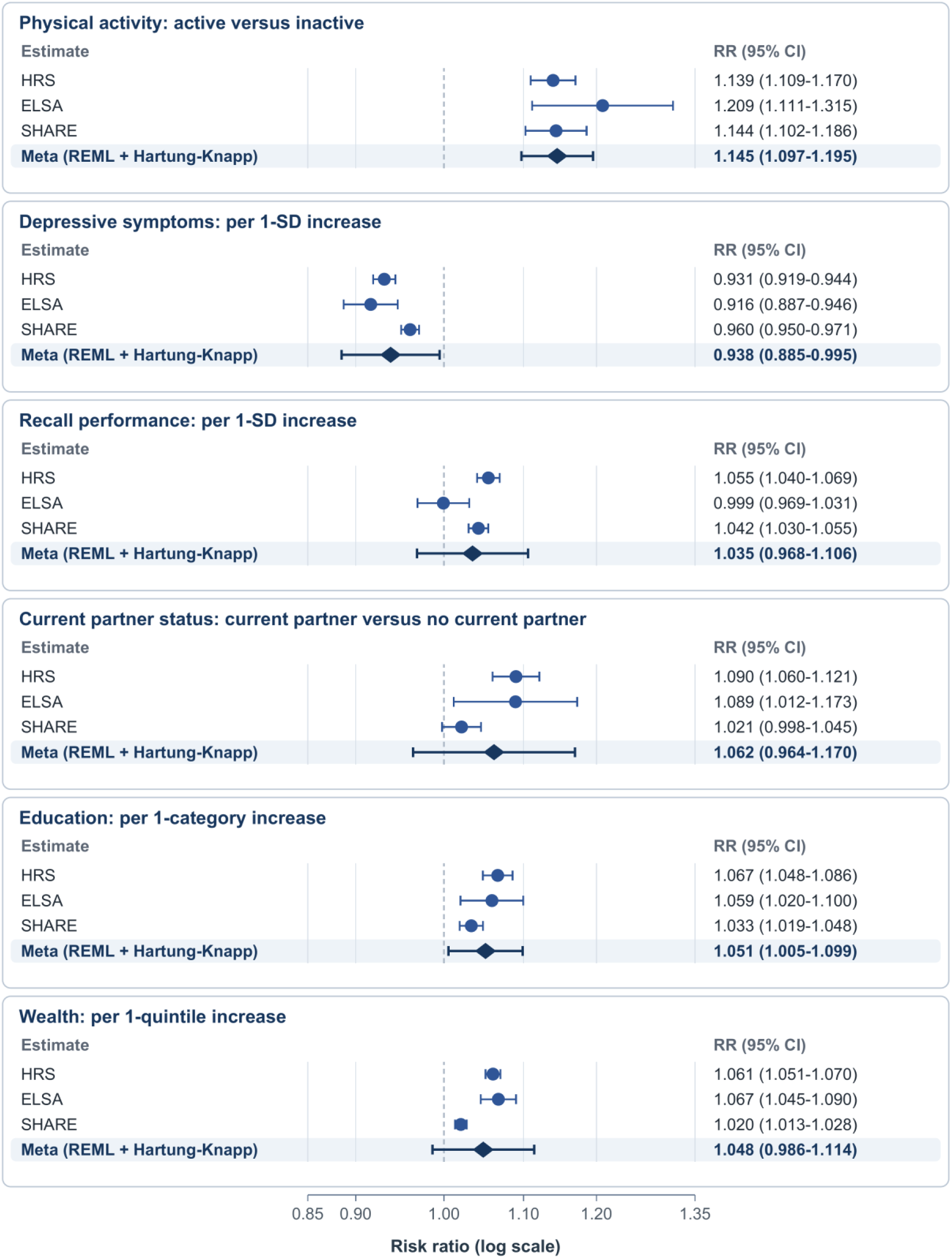
Cohort-Specific and Three-Cohort Meta-analysis Estimates for ADL Maintenance. The figure presents cohort-specific risk ratios and 95% confidence intervals for HRS, ELSA, and SHARE, together with three-cohort random-effects meta-analysis estimates using REML estimation and Hartung-Knapp intervals. CHARLS was not included in the primary meta-analysis. Indicators with high between-cohort heterogeneity should be interpreted cautiously in conjunction with the manuscript text. Abbreviations: ADL, activities of daily living; CI, confidence interval; ELSA, English Longitudinal Study of Ageing; HRS, Health and Retirement Study; REML, restricted maximum likelihood; RR, risk ratio; SHARE, Survey of Health, Ageing and Retirement in Europe. Alt text: Forest plot of risk ratios and 95% confidence intervals for ADL maintenance for six indicators. Within each indicator, rows are shown for HRS, ELSA, SHARE, and the three-cohort random-effects meta-analysis. The horizontal axis is logarithmic with a reference line at a risk ratio of 1.00. CHARLS is not included in the primary meta-analysis.

**Table 3.** Three-Cohort Random-Effects Meta-analysis and Holm-Adjusted Results for ADL Maintenance. Compatible cohort-specific log risk ratios from HRS, ELSA, and SHARE were combined using random-effects meta-analysis. Between-cohort variance was estimated by restricted maximum likelihood, and confidence intervals used the Hartung-Knapp method. The six prespecified two-sided meta-analysis P values formed one confirmatory family and were adjusted using the Holm procedure.

| Indicator | Meta RR | 95% CI | P value | Holm-adjusted P value | I<sup>2</sup> | $\tau^2$ | Q | Q P value | Pooling status |
| --- | --- | --- | --- | --- | --- | --- | --- | --- | --- |
| Physical activity: active vs inactive | 1.145 | 1.097–1.195 | .005 | .032 | 0.6% | 0.000003 | 1.72 | .422 | Pooled |
| Depressive symptoms: per 1-SD increase | 0.938 | 0.885–0.995 | .043 | .201 | 88.0% | 0.000461 | 16.83 | <.001 | Pooled with heterogeneity caution |
| Recall performance: per 1-SD increase | 1.035 | 0.968–1.106 | .157 | .238 | 89.0% | 0.000550 | 9.89 | .007 | Pooled with heterogeneity caution |
| Current partner: yes vs no | 1.062 | 0.964–1.170 | .116 | .238 | 81.7% | 0.001307 | 13.26 | .001 | Pooled with heterogeneity caution |
| Education: per 1-category increase | 1.051 | 1.005–1.099 | .040 | .201 | 70.9% | 0.000258 | 7.91 | .019 | Pooled with heterogeneity caution |
| Wealth: per 1-quintile increase | 1.048 | 0.986–1.114 | .079 | .238 | 95.3% | 0.000565 | 51.86 | <.001 | Pooled with heterogeneity caution |

1. RR indicates risk ratio; CI, confidence interval; ADL, activities of daily living; REML, restricted maximum likelihood.
2. I<sup>2</sup>, $\tau^2$ , and Cochran's Q describe estimated between-cohort heterogeneity. With only three contributing cohorts, heterogeneity measures were interpreted cautiously. Hartung-Knapp confidence intervals were based on two degrees of freedom and were also interpreted cautiously.
3. CHARLS was not included in the primary meta-analysis.
4. The physical activity hypothesis was the only one of the six prespecified hypotheses to meet the Holm-adjusted criterion.
5. The other five hypotheses did not meet the prespecified Holm-adjusted criterion; this does not establish that the corresponding associations were absent.

For depressive symptoms, the pooled RR per 1-SD increase was 0.938 (95% CI, 0.885– 0.995; *P* = .043; Holm-adjusted *P* = .201), with substantial estimated between-cohort heterogeneity (I² = 88.0%). For recall performance, the pooled RR per 1-SD increase was 1.035 (95% CI, 0.968–1.106; *P* = .157; Holm-adjusted *P* = .238), with similarly substantial heterogeneity (I² = 89.0%). Neither hypothesis met the prespecified Holm-adjusted criterion.

The pooled RR for having versus not having a current partner was 1.062 (95% CI, 0.964– 1.170; *P* = .116; Holm-adjusted *P* = .238), with considerable estimated between-cohort heterogeneity (I² = 81.7%). The pooled RR per 1-category increase in education was 1.051 (95% CI, 1.005–1.099; *P* = .040; Holm-adjusted *P* = .201), with appreciable estimated heterogeneity (I² = 70.9%). Neither hypothesis met the prespecified Holm-adjusted criterion.

The pooled RR per 1-quintile increase in wealth was 1.048 (95% CI, 0.986–1.114; *P* = .079; Holm-adjusted *P* = .238), with substantial estimated between-cohort heterogeneity (I² = 95.3%). The wealth hypothesis did not meet the prespecified Holm-adjusted criterion.

### Heterogeneity interpretation

Estimated between-cohort heterogeneity was minimal for physical activity, lower but still appreciable for education and comparatively high for depressive symptoms, recall performance, current partner status and wealth. Pooled estimates were interpreted alongside the corresponding cohort-specific estimates. Because only three cohorts contributed to each meta-analysis, both the heterogeneity estimates and the Hartung–Knapp intervals were interpreted cautiously. Additional heterogeneity assessments and sensitivity analyses are reported in Appendices S6–S7.

## Discussion

### Principal findings

In this harmonised longitudinal analysis of three international ageing cohorts, we examined behavioural, psychological, cognitive, social and socioeconomic indicators in relation to subsequent ADL maintenance among older adults with persistent pain. Physical activity was associated with a higher probability of ADL maintenance in HRS, ELSA and SHARE, and the physical activity hypothesis was the only one of the six prespecified hypotheses to meet the Holm-adjusted criterion in the three-cohort meta-analysis. The other five hypotheses did not meet this criterion, and their estimates should be interpreted in light of varying cohort-specific patterns and generally appreciable between-cohort heterogeneity. Failure to meet the prespecified Holm-adjusted criterion does not establish that the corresponding associations were absent; conversely, these observational findings do not establish causality or show that modifying any single indicator would preserve ADL function.

### Clinical implications

Assessing functional ability alongside pain symptoms is consistent with multidimensional pain assessment in older people [25]. Physical activity is measurable and potentially modifiable [26], and previous prospective work linked lower activity to later disability in older adults with chronic pain [27]. Although observational findings do not establish benefit from increasing activity, the consistent cohort-specific estimates and Holm-adjusted result support evaluating tailored activity-support strategies.

### Public health implications

Persistent pain and functional loss affect health and long-term-care needs [1,2]. Public health approaches should consider functional maintenance alongside symptom burden and conditions enabling activity and independence. Although this study did not evaluate an intervention, it supports testing equitable, safe and contextually appropriate activity-support strategies, including how pain, mobility, access and socioeconomic circumstances affect feasibility and uptake.

### Interpretation of other indicators

Higher depressive-symptom scores were associated with a lower probability of ADL maintenance in all three cohorts, but the pooled estimate showed substantial between-cohort heterogeneity and the depressive-symptoms hypothesis did not meet the prespecified Holm-adjusted criterion. Recall-performance estimates were above 1.00 in HRS and SHARE but close to 1.00 in ELSA; the pooled estimate was also heterogeneous, and the recall-performance hypothesis likewise did not meet this criterion. Although memory and related cognitive abilities are relevant to everyday functioning in later life [28], the present psychological and cognitive findings were less consistent and did not provide the same confirmatory evidence as the physical activity result.

Current-partner estimates were above 1.00 in all three cohorts but were smaller and closer to 1.00 in SHARE. Although broader social vulnerability measures have been associated with functional decline, they are not directly equivalent to current partner status [11]. Education and wealth estimates were also above 1.00 in all three cohorts, but neither hypothesis met the prespecified Holm-adjusted criterion. Differences in social context, welfare systems, measurement, relative socioeconomic position and sample composition may help explain the cross-cohort variation. Failure to meet this criterion should not be interpreted as evidence that these associations were absent.

### Strengths

This study drew on three major ageing cohorts and applied prespecified harmonised operational definitions of persistent pain, six resource-related indicators and ADL maintenance. Its longitudinal design separated persistent-pain ascertainment, baseline indicator measurement and subsequent assessment of ADL maintenance. The six hypotheses and their confirmatory multiplicity family were prespecified before the primary analyses.

Cohort-specific modified Poisson estimates were presented alongside random-effects pooled estimates, preserving visibility of cross-cohort differences. Multiplicity was addressed using the Holm procedure, and robust variance estimation accounted for repeated HRS person-windows. Indicator-specific complete-case samples avoided requiring complete data on all six indicators for every analysis.

## Limitations

Because this was an observational study, residual confounding and reverse causation cannot be ruled out, and the estimated associations should not be interpreted causally. Although cohort-specific measures were harmonised to common operational definitions, the underlying assessments were not identical, and several pooled associations showed substantial estimated between-cohort heterogeneity. Differences in measurement, population composition and contextual conditions may help explain this variation, but these possibilities could not be distinguished with only three cohorts. Both the heterogeneity estimates and the corresponding pooled associations should therefore be interpreted cautiously [23].

Each indicator was analysed in a separate complete-case sample, so analytic sample composition varied and missing data may have introduced selection bias. Complete-case analysis and loss to follow-up can introduce bias under some missing-data mechanisms [29,30]. Sampling weights, inverse-probability weighting for T2 retention and multiple imputation were not used; population representativeness may therefore be limited, and selection related to attrition or missingness cannot be ruled out. Because death and other non-observed T2 states were not classified as ADL-maintenance failure, interpretation is limited to observations with valid follow-up ADL data. HRS used a person-window estimand, whereas ELSA and SHARE used person-level analyses, which may limit exact cross-cohort comparability. Finally, the harmonised physical-activity indicator did not identify the activity type, dose or delivery strategy that would be appropriate or effective for older adults with persistent pain.

## Conclusion

Among older adults with persistent pain, physical activity was associated with a higher probability of subsequent ADL maintenance in all three cohorts, and the physical activity hypothesis was the only one of the six prespecified hypotheses to meet the Holm-adjusted criterion in the three-cohort meta-analysis. The other five hypotheses did not meet this criterion, which should not be interpreted as evidence that the corresponding associations were absent. The findings highlight the relevance of considering ADL function alongside pain symptoms in pain research and clinical assessment. However, the observational evidence does not establish that increasing physical activity would preserve ADL independence or that any specific activity programme would be effective.

## Author Contributions

Lei Zhang conceived the study, developed the analytic methodology, curated and analysed the data, prepared the tables and figures, and drafted the manuscript. Wenjing Zhang and Linchao Li contributed to methodology, validation of the analytical workflow and interpretation of the results. Yuzhu Ye, Xiaona Zhu, Liang Shao, Huidi Yang, Yue Hu and Yuheng Li contributed to the literature review, interpretation of the findings and critical revision of the manuscript. Hai Lin and Wujun Geng jointly supervised the study, contributed to study conception, methodology and interpretation, and critically revised the manuscript. Wujun Geng provided overall project administration. All authors reviewed and approved the final manuscript and agree to be accountable for the work.

## Data Availability

The de-identified data analysed in this study are available to eligible researchers from the Health and Retirement Study, the English Longitudinal Study of Ageing and the Survey of Health, Ageing and Retirement in Europe through their respective official registration and data-access procedures. Access is subject to the terms and conditions of each cohort. The authors are not authorised to redistribute the person-level data. No new participant-level data were generated for this study.

## AI Use Disclosure

OpenAI ChatGPT and Codex were used for language editing and formatting adjustment. These tools did not determine the research question, analysis definitions, statistical specifications, scientific interpretation or conclusions. All AI-assisted code, processing outputs, statistical results, references and text were reviewed and verified by the authors, who take full responsibility for the accuracy and integrity of the submitted work. No AI system is listed as an author.

## Supplementary Data

Appendix S1. Supplementary methods

Appendix S2. Cohort harmonisation and variable definitions (Tables S1–S4)

Appendix S3. Participant flow and indicator-specific analytic samples (Tables S5–S6)

Appendix S4. Model diagnostics and reproducibility checks (Table S7)

Appendix S5. Non-estimability and audit of fixed four-year follow-up duration in the three primary cohorts (Table S8)

Appendix S6. Between-cohort heterogeneity assessment (Tables S9–S10)

Appendix S7. Completed sensitivity analyses (Tables S11–S12)

Appendix S8. Role of CHARLS as a candidate extended sensitivity cohort

Appendix S9. Analyses not performed

## References

1. Fayaz A, Croft P, Langford RM et al. Prevalence of chronic pain in the UK: a systematic review and meta-analysis of population studies. BMJ Open 2016;6(6):e010364. 10.1136/bmjopen-2015-010364.

2. Beard JR, Officer A, Carvalho IA de et al. The World report on ageing and health: a policy framework for healthy ageing. The Lancet 2016;387(10033):2145–54. 10.1016/s0140-6736(15)00516-4.

3. Kawashima A, Komatsu A, Jin X et al. Chronic pain and decline in activities of daily living among community-dwelling older adults: a systematic review of longitudinal studies. Eur Geriatr Med 2025;16(6):2085–96. 10.1007/s41999-025-01299-5.

4. Cheng F, Wang Z, Li Y et al. Longitudinal associations of chronic pain severity trajectories and number of pain site trajectories with risk of limitations in ability in daily activities: Evidence from two 10-year prospective cohort studies. J Nutr Health Aging 2026;30(3):100781. 10.1016/j.jnha.2026.100781.

5. Whitson HE, Duan-Porter W, Schmader KE et al. Physical Resilience in Older Adults: Systematic Review and Development of an Emerging Construct. J Gerontol A Biol Sci Med Sci 2016;71(4):489–95. 10.1093/gerona/glv202.

6. Geneen LJ, Moore RA, Clarke C et al. Physical activity and exercise for chronic pain in adults: an overview of Cochrane Reviews. Cochrane Database Syst Rev 2017;4:CD011279. 10.1002/14651858.cd011279.pub3.

7. Leung DKY, Fong APC, Wong FHC et al. Nonpharmacological Interventions for Chronic Pain in Older Adults: A Systematic Review and Meta-Analysis. Gerontologist 2024;64(6):gnae010. 10.1093/geront/gnae010.

8. Cunningham C, O’ Sullivan R, Caserotti P et al. Consequences of physical inactivity in older adults: A systematic review of reviews and meta-analyses. Scand J Med Sci Sports 2020;30(5):816–27. 10.1111/sms.13616.

9. Zhu X, Wang Y, Luo Y et al. Bidirectional, longitudinal associations between depressive symptoms and IADL/ADL disability in older adults in China: a national cohort study. BMC Geriatr 2024;24(1):659. 10.1186/s12877-024-05248-y.

10. Raimo S, Maggi G, Ilardi CR et al. The relation between cognitive functioning and activities of daily living in normal aging, mild cognitive impairment, and dementia: a meta-analysis. Neurol Sci 2024;45(6):2427–43. 10.1007/s10072-024-07366-2.

11. Cappelli M, Bordonali A, Giannotti C et al. Social vulnerability underlying disability amongst older adults: A systematic review. Eur J Clin Invest 2020;50(6):e13239. 10.1111/eci.13239.

12. Depp CA, Jeste DV. Definitions and predictors of successful aging: a comprehensive review of larger quantitative studies. Am J Geriatr Psychiatry 2006;14(1):6–20. 10.1097/01.jgp.0000192501.03069.bc.

13. Li X, Gao L, Qiu Y et al. Social frailty as a predictor of adverse outcomes among older adults: a systematic review and meta-analysis. Aging Clin Exp Res 2023;35(7):1417–28. 10.1007/s40520-023-02421-y.

14. Saadeh M, Welmer AK, Dekhtyar S et al. The Role of Psychological and Social Well-being on Physical Function Trajectories in Older Adults. J Gerontol A Biol Sci Med Sci 2020;75(8):1579–85. 10.1093/gerona/glaa114.

15. Wahrendorf M, Reinhardt JD, Siegrist J. Relationships of disability with age among adults aged 50 to 85: evidence from the United States, England and continental europe. PLoS One 2013;8(8):e71893. 10.1371/journal.pone.0071893.

16. Wu YT, Daskalopoulou C, Muniz Terrera G et al. Education and wealth inequalities in healthy ageing in eight harmonised cohorts in the ATHLOS consortium: a population-based study. Lancet Public Health 2020;5(7):e386–94. 10.1016/S2468-2667(20)30077-3.

17. Gjonça E, Tabassum F, Breeze E. Socioeconomic differences in physical disability at older age. J Epidemiol Community Health 2009;63(11):928–35. 10.1136/jech.2008.082776.

18. Sonnega A, Faul JD, Ofstedal MB et al. Cohort Profile: the Health and Retirement Study (HRS). Int J Epidemiol 2014;43(2):576–85. 10.1093/ije/dyu067.

19. Steptoe A, Breeze E, Banks J et al. Cohort profile: The english longitudinal study of ageing. Int J Epidemiol 2013;42(6):1640–8. 10.1093/ije/dys168.

20. Börsch-Supan A, Brandt M, Hunkler C et al. Data Resource Profile: the Survey of Health, Ageing and Retirement in Europe (SHARE). Int J Epidemiol 2013;42(4):992–1001. 10.1093/ije/dyt088.

21. Zou G. A modified poisson regression approach to prospective studies with binary data. Am J Epidemiol 2004;159(7):702–6. 10.1093/aje/kwh090.

22. Knapp G, Hartung J. Improved tests for a random effects meta-regression with a single covariate. Stat Med 2003;22(17):2693–710. 10.1002/sim.1482.

23. Higgins JPT, Thompson SG, Deeks JJ et al. Measuring inconsistency in meta-analyses. BMJ 2003;327(7414):557–60. 10.1136/bmj.327.7414.557.

24. Holm S. A Simple Sequentially Rejective Multiple Test Procedure. Scandinavian Journal of Statistics 1979;6(2):65–70. 10.2307/4615733.

25. Schofield P, Dunham M, Martin D et al. Evidence-based clinical practice guidelines on the management of pain in older people - a summary report. Br J Pain 2022;16(1):6–13. 10.1177/2049463720976155.

26. Piercy KL, Troiano RP, Ballard RM et al. The Physical Activity Guidelines for Americans. JAMA 2018;320(19):2020–8. 10.1001/jama.2018.14854.

27. Makino K, Lee S, Lee S et al. Daily Physical Activity and Functional Disability Incidence in Community-Dwelling Older Adults with Chronic Pain: A Prospective Cohort Study. Pain Med 2019;20(9):1702–10. 10.1093/pm/pny263.

28. Deary IJ, Corley J, Gow AJ et al. Age-associated cognitive decline. Br Med Bull 2009;92(1):135–52. 10.1093/bmb/ldp033.

29. Sterne JAC, White IR, Carlin JB et al. Multiple imputation for missing data in epidemiological and clinical research: potential and pitfalls. BMJ (Clinical research ed.) 2009;338:b2393. 10.1136/bmj.b2393.

30. Howe CJ, Cole SR, Lau B et al. Selection Bias Due to Loss to Follow Up in Cohort Studies. Epidemiology (Cambridge, Mass.) 2016;27(1):91–7. 10.1097/EDE.0000000000000409.

